# AMPLIFICATION OF THE POWER OF NETWORK HUBS AND DEGREE SKEWNESS OVER INFECTIOUS DISEASE SPREAD DURING LULLS

**DOI:** 10.1101/2025.03.27.25324820

**Authors:** Benjamin Cornwell, Shiyu Ji, Shane G. Henderson, Gen Meredith

## Abstract

Just as individuals’ personal social network connections shape their susceptibility to disease, the structure of larger networks in communities shapes the extent of disease spread. We examine how heterogeneity in network structure at the population level affects the spread of disease – namely, COVID-19 – considering varying levels of disease transmissibility and in-person contact rates. Using dynamic simulations that take into account network structure, social contact rates given contextual features of the community (informed by real-life data on family local family structure, schools, workplaces, and daily shopping activities), and disease infection rates, we first confirm that the presence of network hubs and high network degree skewness results in a higher level of infected peak prevalence with infectious diseases such as COVID-19 during periods of low to moderate transmissibility. However, this effect is amplified during lulls in disease spread and is suppressed during periods of greater transmissibility, rendering social network structure more significant during lulls. Moreover, in the case of already highly transmissible diseases, the role of hubs and severe degree skewness is already more continually suppressed.

## INTRODUCTION

The link between social networks and health is the basis of one of the liveliest topics of research in sociology. There are numerous influential studies on this empirical connection [1, 2, 3, 4]. There are many avenues of research in this vein, including such specific issues as the link between levels of social connectedness (conceptualized in various ways) and the common cold, functional health, cognitive health, and mortality. Much of this work focuses on individual-level behaviors and the functions and effects of their social ties, including social support, coping, access to resources and social influence [5, 6, 7, 8, 9, 10, 11, 12, 13, 14].

Related work elevates the focus to community-level risks and outcomes, such as the epidemic spread of disease and attendant public health policies [2, 15]. This body of work leads to the general conclusions that individuals’ health depends not just on their individual connections, but also the connections that exist around them, in the broader community.

Along these lines, this paper focuses on a particular health threat, which is the presence of rapidly spreading viruses that lead to the epidemic spread of deadly diseases, such as the human immunodeficiency virus (HIV), Influenza, the respiratory syncytial virus (RSV), and severe acute respiratory syndrome coronavirus 2 (SARS-CoV-2). Given the recent spate of such disease epidemics around the world (some of them ongoing or recurring), social scientists have also sought to understand the factors affecting their genesis, partly in an effort to inform prevention strategies, and in an urgent effort to reduce morbidity and mortality. Consistent with research on other viruses, key factors that influence spread include local climate and the transmissibility of the virus itself [16, 17]. Importantly, social scientists have focused increasingly on the effects of social-structural factors on spread, such as population density, levels of social and material disadvantage, racial/ethnic composition, public transportation systems, vaccination rates, and politics, among others [18, 19, 20, 21, 22, 23, 24].

While understanding broad epidemiological factors influencing viral spread is crucial, considering the granular details of interpersonal connections is equally important. It has been demonstrated that social networks significantly impact disease transmission dynamics. An individual’s level of connectedness to others – especially the number of their connections and their level of contact with others in everyday settings – have proved to be some of the most important factors that contribute to individual susceptibility and, by extension, spread of disease within communities.

In this paper, we expand on this work by considering the role that *larger social network structure* plays in the spread of disease – and thus the risk of larger epidemics and pandemics. We do so by considering the interplay of two key sets of factors: (1) The characteristics of the social networks that exist within communities; and (2) The transmissibility of a given virus within a community, given its inherent infectiousness. We pay particular attention to the possibility that social network structure conditions the role that infectiousness plays in the risk of the epidemic spread of disease throughout the community.

### Influence of Network Hubs and Degree Skewness on the Spread of Disease

It is well established in the social-epidemiology literature that there is considerable heterogeneity with respect to network degree [25, 26]. In other words, most social networks are characterized by a skewed degree distribution that corresponds to a network with a few highly connected people known as “hubs” and many people with fewer ties. The same can be said for the distribution of the number of social contacts people have daily – with some people (e.g., grocery store clerks, teachers) having numerous contacts and others having few by comparison. Well-connected individuals often play larger roles in spreading diseases like HIV/AIDS, where those occupying bridging positions in sexual networks significantly influenced the epidemic [27, 28].

Reflecting the variation among individuals, different populations or communities have different network structures connecting people [29, 30]. Studies have suggested that greater heterogeneity in network connectedness, as opposed to the presence of uniformly densely connected or random networks, have contributed to the epidemic spread of the disease at the population level [31, 32, 33, 34].

Recent epidemics and pandemics over the last few decades – including those relating to HIV/AIDS and COVID-19 – have underscored the importance of understanding the role of social networks and network degree distribution in disease transmission. Research – including analysis of contact tracing data – shows that people with more social connections are more likely to contract and transmit SARS-CoV-2, and that large gatherings facilitate epidemic spread [24, 27, 33, 34, 35]. This insight underpins disease prevention and control strategies like social distancing, community quarantines, and university lockdowns to curb COVID-19 and other pandemics via reducing person-to-person contact [36, 37, 38, 39, 40, 41, 42, 43], since the involvement in social activities and sustained social contact [46] is crucial for virus transmission, whether through respiratory fluids or surfaces.

At network level, past studies tend to show a highly skewed distribution in the transmission network of COVID-19 network [37, 44]. It has been reported that the maximum degree in the transmission network in one wave ranges from 64 to 127, while the average degree in the transmission network is just between 3.8 and 6.4 [45].

Although the spreading network differs from a potential contact network, the above results suggest that a few individuals act as hubs in their community networks, with this social network heterogeneity potentially contributing to virus “hot spots” [29, 46, 47, 48]. Despite this, the specific impact of a contact network’s degree distribution on disease spread – and its interaction with transmission rate – remains unclear. This uncertainty is partly due to the lack of empirical data on contact networks, which complicates efforts to accurately model transmission dynamics.

Additionally, the transmissibility of a virus varies widely across virus types and their variants. The initial Alpha variant of SARS-CoV-2, for example, had an estimated basic reproduction number (R0) of around 2.9 [49], while the Delta variant’s R0 is estimated between 3.2 and 6 [50, 51]. The more recent Omicron XBB.1.5 variant is thought to have much higher transmissibility, with a reproducibility estimate that might exceed 20, though this estimate seems to depend on local context, including vaccination rates and related factors [52]. This is on par with more contagious diseases such as measles, which is often pinned at the 12-18 range. [52].

Given the variation in transmission rates across different types and variants of viruses, it remains uncertain whether social network degree skewness – typically associated with increased epidemic spread in scenarios involving superspreaders, or a more skewed social network connectedness (i.e., “degree”) distribution – continues to play a significant role when transmission rates are extremely high. The gap in empirical data for contact network makes the question particularly well-suited to simulation modeling, which could help assess the influence of degree skewness under conditions of high transmissibility.

Seeing these challenges, this paper expands on existing research by exploring the role of social network structure in disease spread, using simulations with SARS-CoV-2-related parameters as a recent example. It investigates whether, and to what extent, heterogeneity within individuals’ social networks, as described above, shapes the prospect of epidemic spread across variants that have different levels of transmissibility. Disease transmission hinges on social connectedness, so it is vital to understand how the structure of a network in a given community might shape its susceptibility to an epidemic of varying speed. To effectively explore the role of the hub, we conducted simulations informed by empirical data, and isolate the effect of the heterogeneity in individual contacts, while holding constant the total contact in a network.

## MATERIALS AND METHODS

### Social Network Structure Used for Simulations

We first describe the construction of random social networks for simulating the epidemic. In this study, network generation is decomposed into two steps. We initially construct a random base social network to represent everyday social interactions based on families, schools, workplaces, and daily shopping activities, informed by real-life data (see methodological details in the Appendix). These various venues, termed social foci, serve as focal points where individuals regularly engage in social interactions and daily activities [53]. The construction process generates basic family structures, assigns adults to workplaces and children to schools, and assigns edge weights – approximations of the strength of the contacts – to represent contact levels, assuming uniform random interaction within social foci. The edge weight between two individuals sharing the same foci is inversely proportional to the size of the respective social foci. For example, if a school consists of N students, the edge weight between any two random students in the school is 1/N. If two individuals share more than one foci, the edge weight between them is the accumulation of the weights brought by each of the focus.

For each randomly simulated base network, we introduce heterogeneity in degree distribution by adding friendship ties, which can be assortative (yielding a highly skewed distribution) or disassortative (yielding a centralized distribution), aiming for an average total contact level 50% above the base network. This process is referred to as the preferential attachment process in the following text. Friendship edges are added until the target average is reached, with edge weights chosen uniformly at random from the base network, resulting in networks with varying degree distributions but identical total contact levels. While the spreading network of COVID-19 is estimated to be highly skewed, empirical and theoretical work on the degree distributions of individuals’ daily connection networks yields [21, 54, 55, 56] varied estimates– some suggesting a highly skewed, scale-free distribution [57, 59], others assuming more bell-shaped distributions but with an underlying small-world signature [58]. To acknowledge this difference, we manipulate a single skewness parameter to adjust the edges and examine 12 skewness parameters (α), ranging from -4 to 4, to investigate their impact on degree distribution and its interaction with disease transmissibility. With a skewness parameter greater than 0, the process emulates a preferential attachment mechanism [59], wherein nodes with more connections are more likely to acquire new links. For each base network simulation, we create 12 networks with varying degrees of skewness, maintaining consistent total contact levels across them. The process of creating social networks with specific degree distributions is detailed in the Appendix.

### Comparison with Existing Models

Our model effectively balances the scale-free structure observed in real-world social networks – capturing dyadic interactions, such as friendships that carry infection risks from specific contacts – with broader community-level activities like schooling and shopping, where infection risks arise from shared environments rather than specific individual connections. In constructing the foci network, we incorporate key factors like contact frequency, social-foci size, and exposure duration across settings such as households, workplaces, grocery stores, and schools. This enables our model to more accurately represent variations in transmission potential.

Unlike the Stochastic Block Model which restricts each individual to a single group, our approach allows individuals to participate in multiple groups, reflecting the overlapping social interactions common in real life [60]. While preferential attachment models emphasize high-degree centrality, our method integrates context-sensitive exposure patterns within different social foci [58]. Additionally, classic Erdős-Rényi models [61], which do not take social role differentiation into account, are less suited to capturing the nuanced transmission dynamics that our model achieves by considering both contact intensity and transmission risk.

#### Procedure for Generating the Household Network

To construct a network that meaningfully encapsulates the structures and contexts that characterize daily social life, we employ a social-foci framework as our foundational model. This concept, introduced by Feld [62], identifies various venues – ranging from workplaces and schools to churches, bars, and shopping centers – as focal points where individuals regularly engage in social interactions and daily activities. As an initial step in our network construction, we generate a basic family structure within this broader social-foci framework. To accomplish this, we iterate through individuals within our simulated sample in a sequential manner. At each step, there is an 80% probability that a new individual is marked as part of a household unit, paired with the subsequent individual in the sequence. Once this pairing is established, zero to three subsequent individuals are randomly selected via uniform random sampling to represent their children. After completing the assembly of one household, we reset the procedure and apply the aforementioned steps to generate subsequent households, beginning with the next available individual in the sample.

#### Procedure for Generating Social Network Foci

In addition to families, we integrate individuals into other distinct social foci. For the purpose of this study, we concentrate on three primary social foci: workplaces and grocery stores for adults and schools for children. These social-structural assumptions are not so much intended to imitate the real-life scenario, except that individuals are bound into social foci, wherein much of their contact occurs [54]. As a result, during the foci-network construction, the sizes of these foci and the selection of parameter estimates approximate the density of real cities, but they are not the variable of interests.

To model *workplaces*, we consider the total network size N and generate 18N/1000 workplaces, meaning that there are 18 workplaces for every 1,000 individuals in the network, which is 180 workplaces in our network since N = 10,000. The U.S. population in 2023 was around 335.0 million [63], and, as of 2020 [64], there are 6.1 million employer firms in the United States [65]. This is roughly 18 employer firms per 1000 individuals. Sixty percent of adults in the network are designated as employed. Each employed adult is then randomly assigned to a workplace, following a power-law distribution with an exponent of -1/2. In particular, for workplace number *k* (*k ∈* {1,2,…, 180}), the probability that an employed adult being assigned to each of the workplace is proportional to 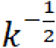.

For *schools*, we posit that one educational institution exists for every 2,500 individuals in the network. According to the United States Bureau of Labor Statistics, the employment rate as of May 2023 is 60.2%. Each child is given a 75% probability of being enrolled in a school. According to National Center for Education Statistics [66], there are 50.8 million students enrolled in public PK-12 education and 4.7 million students enrolled in private PK-12 education. According to the United States Department of Justice, Office of Juvenile Justice and Delinquency Prevention [67], 72.8 million Americans are between the ages of 0 and 17. This is an enrollment rate of 76.2%. School assignments for children are made through uniform random sampling across the available schools.

For *grocery stores and shopping*, similar to schools, we assume the existence of one grocery store for every 2,500 individuals. According to the United States Department of Agriculture, Economic Research Service [68], there are 115,526 food stores in the United States, which is an average of one out of every 2,900 individuals. Within each household, an adult is randomly selected to undertake grocery shopping responsibilities. It is assumed that this individual frequents the same grocery store, chosen via uniform random sampling from the available options.

#### Determining Contact Level per Household and Foci

Within these foci, we assume individuals to interact with each other uniformly at random, as epidemiologists typically assume of the entire population. The strength of these contact ties, which represents frequency of contact, is assumed to be inversely proportional to the size of the foci which entails them. To put this assumption in context, for instance, a member in a workplace of six workers has a higher average contact level with every individual co-worker compared to a worker in a workplace of 100.

In addition, people spend much less time on average on grocery shopping each week than on workplaces, families and schools. We assume that the chance of being infected by disease is lower for each possible contact happening in grocery stores by a factor of *k*. Here we set k as 1/40, which comes from the assumption that an individual works for 40 hours on average per week, while this individual goes to the grocery store for one hour in total weekly.

Mathematically, the contact level *w*_*i,j*_ between individual *i* and *j* is the sum of the contacts that they have in each of their shared foci *g*. That is,

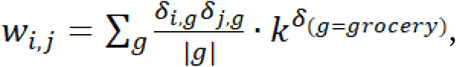

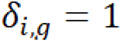 when individual belongs to the *g* foci and 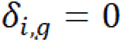 if otherwise.| *g* | is the number of individuals included in foci *g*. *δ*_(*g* = *grocery*)_ = 1 when the foci is a grocery store, under which the chance of infected per unit contact is discounted by *k*.

### Generating a Degree-Skewed Social Network Given a Base Network

Based on the foci network, we now describe the key independent structural variable, which generates these more- or less-skewed networks. We introduce this variable by adding friendship ties, generated independently from the social foci structure of our imaginary city. The variation comes in how assortative or disassortative these network ties are. In particular, we start by setting a target average total contact, at 50% above the base network. We define total contact as \*sum* – {*e*\*in*{*E*}}*w*(*e*), where *E* denotes the set of all edges in the graph, and *w*(*e*) is the weight of the edge.

We then add edges sequentially until this target is reached. We choose the first node of an edge uniformly at random from all nodes, and choose the partner node, say *i*, with probability proportional to *d*(*i*)^α^, where *α* encodes the importance of degree in deciding the partner. When *α* = 0, partners are chosen uniformly at random. When *α* > 0, partners are chosen with higher probability if they have a higher degree. When *α* > 0, partners are chosen with lower probability if they have higher degree. We used the following skewness parameters: -4, -2, 0 (meaning no skewness compared to the base network), 1, 1.5, 2, 2.5, 3, 3.2, 3.5, 3.8 and 4. The weight of the edge is chosen uniformly at random from the existing edges in the base network. Edges are added until the target average is reached. Networks generated in this way have different degree distributions, but the same total contact.

To illustrate the effectiveness of our process on altering the network degree distribution, we sample a social foci base network comprising 10,000 individuals. The resulting base network has an initial total contact level (i.e. the sum of the existing edge weights) of 8,251.0. After each preferential attachment process with varying skewness parameters, every final network reaches a final contact level of 12376.8. The degree distributions of the exemplar final networks could be found in Figure 1. In Figure 1, for a network adjusted with a skewness parameter of -4, the individual with the maximum weighted degree has a contact level of 4.13, while the individual with the minimum weighted degree has a contact level of 1.49. Conversely, in a network adjusted with a skewness parameter of 4, the maximum individual contact level skyrockets to 1,444, while the minimum drops to .63 (right tail of the distribution that has skewness equal to 4 is omitted from Figure 1 due to space restraints.).

**Fig 1.**
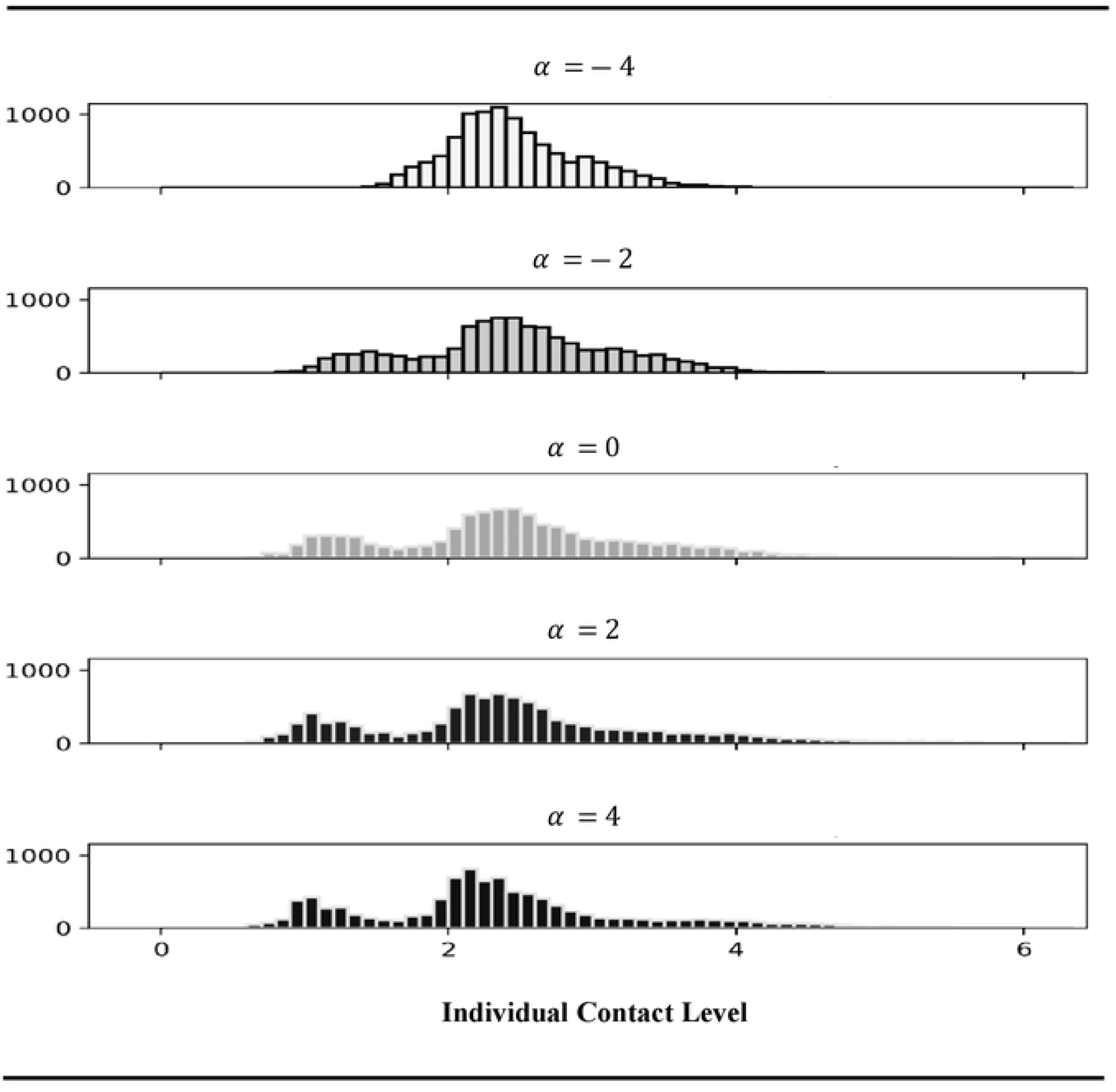
The Simulated Impact of *α* on the Skewness of a Social Context-Level Distribution (N = 10,000) *Note:* This figure is based on a comparison of the aggregate weighted degree distributions among individuals in a sample of the final networks that are based on one of the randomly-generated base foci-networks we examine in this paper. Alpha parameterizes the amount of assortativity in the ad-hoc friendships which are added to the same base network of schools, work, family, and grocery social circles. Total contact in the society is equal across networks, and thus the mean level of contact per person is constant. The long right tails are omitted for better visualization.

In Figures 2 and 3, we compare two randomly generated networks based on the same base network with 10,000 individuals: one with a highly skewed degree distribution (*α* = 4, Figure 2) and the other with a more centered distribution (*α* = −4, Figure 3). To facilitate better visualization in both figures, we only plot 500 nodes, selecting the 10 with the highest node degree in the graph and randomly choosing the other 490. As shown in the visualizations, the highly skewed network (Figure 2) features a single hub with connections to most of the other nodes, indicating a centralized structure. In contrast, the disassortative network (Figure 3) lacks a node with an extremely high degree, suggesting a more evenly distributed network without dominant hubs.

**Fig 2.**
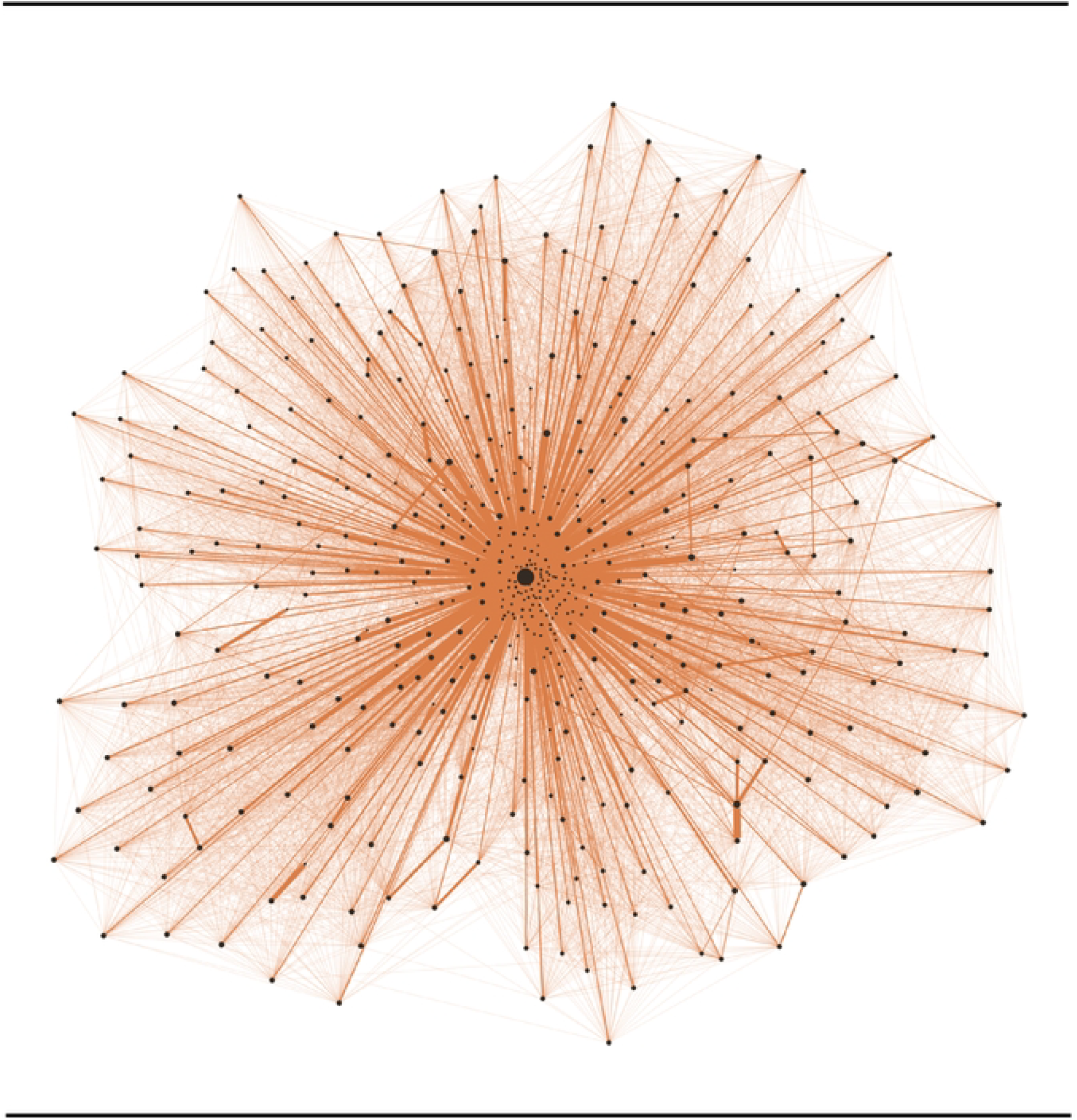
Focused Illustration of a Small Part (500 Nodes) of the Social Network with the Presence of Hubs/Skewness in the Community Infection Simulation. *Note:* The thickness of network edges represents the amount of total contact between individuals, and the size of the node is the contact level of the individual. To generate this graph, we choose the top 10 individuals with the highest contact level and then randomly sample 490 other individuals in the graph. We can see that the focal person in the middle is very well-connected, while other individuals are significantly less well-connected, which exemplifies a larger trend across this degree-agnostic network.

**Fig 3.**
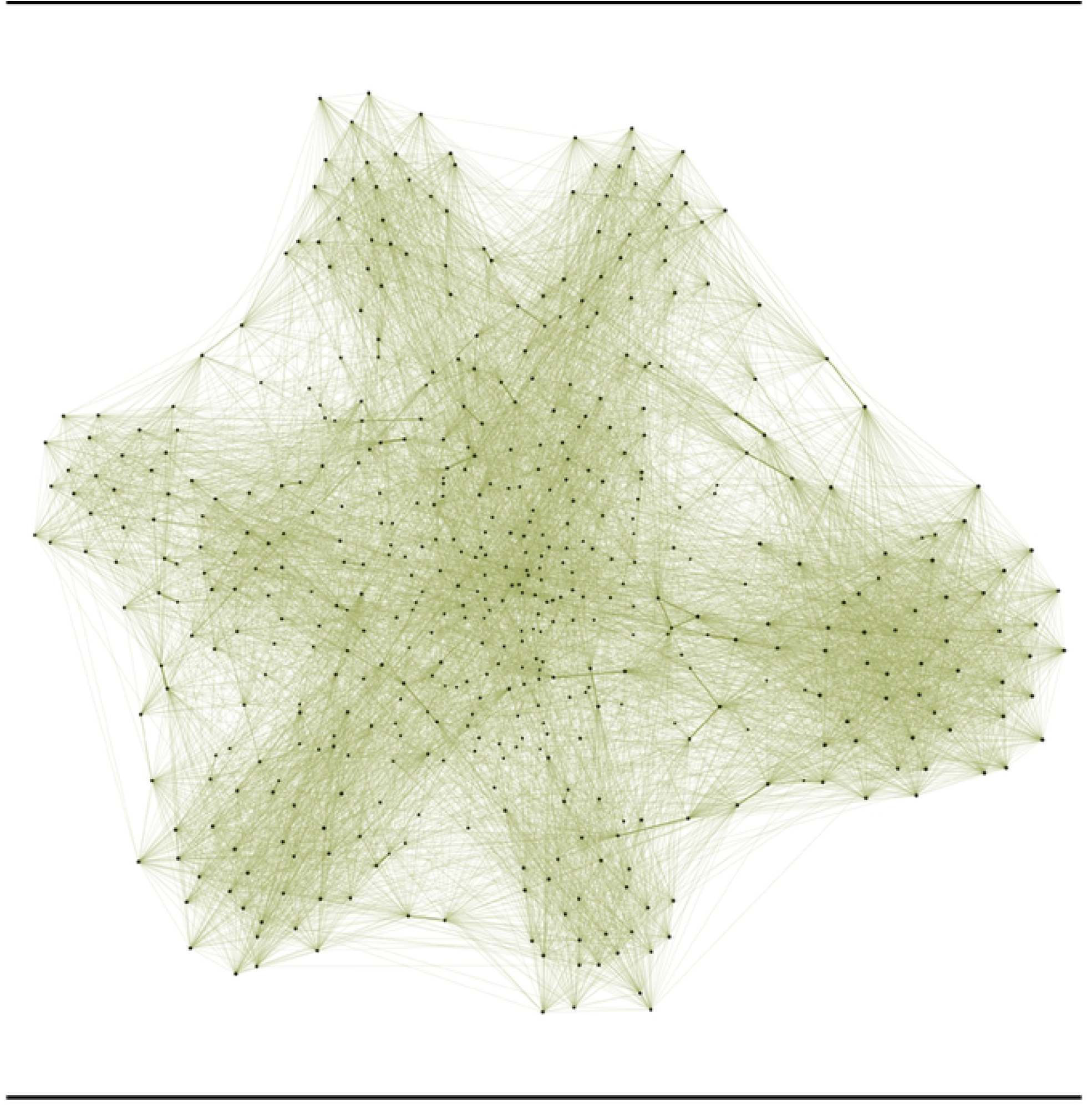
Focused Illustration of a Small Part (500 Nodes) of the Social Network with the Lowest Prevalence of Hubs/Skewness in the Community Infection Simulation. *Note:* The thickness of network edges represents the amount of total contact between individuals, and the size of the node is the contact level of the individual. To generate this graph, we choose the top 10 individuals with the highest contact level and then randomly sample 490 other individuals in the graph. We can see that every node seems to be evenly connected, which exemplifies a larger trend across this degree-agnostic friendship network.

### Simulating Epidemic Spread and Population-Level Susceptibility

To simulate the spread of a disease on each of the generated networks, we employ a continuous-time probabilistic SEIR (susceptible, exposed, infected, recovered) model. At the start of the simulation, 5% of individuals are randomly selected to be in the infected state (positive for COVID-19). Then, based on their weighted contact with others in the network, they expose others to the disease. Transitions between states occur after a time which is exponentially distributed, which is a common assumption in epidemic modeling due to the memoryless property that simplifies the process by treating transition rates as constant over time [69^69^]. Exposure is moderated by the total contact an individual has with infected individuals, while the transition rates from Exposed to Infected and from Infected to Recovered are constant across the population. However, we also acknowledge that transitions may not always follow an exponential distribution, and this will leave to future work to discover [69]. All transition rates are constant over time. We used EoN python package [70], which adopts the Gillespie algorithm, to efficiently simulate the continuous-time Markov chain which this process describes. For better clarity and flow, we detail the experiment-specific, full simulation replication settings at the beginning of the results section

Transition rates between states are chosen to approximate the early spread of COVID-19 [71]. For COVID-19, there is conflicting evidence on the rates of re-susceptibility (*β*_*rs*_). Guedes et al. [72] measured reinfections amongst healthcare workers March 2020 to March 2022, and found that reinfection was present in only 5% of cases, with an average reinfection interval of 429 days. Likewise, Hu and Geng [73] used a heterogeneous SIRS model to estimate clusters of parameters across the United States in April 2020, finding a 95% highest probability range of the rate of re-infection *β*_*rs*_ of (1.181 × 10^−7^,0.0047). The upper value represents an average reinfection time of 213 days. On the other hand, Nguyen et. al [63] analyzed 17 studies and reported an average re-infection interval of 178.9 days, while the variation between studies is large. In addition, they mentioned that the reinfection rates are higher with the Omicron Variants, which partially explains the inconsistencies across studies. In our study, focusing on the total number of infections in a single outbreak, we set our reinfection rate to zero to clearly observe the effect. However, to assess the peak of the outbreak (i.e., the maximum number of individuals being infected at the same time), we conducted robustness checks with two other reinfection rates: one akin to influenza, resulting in reinfection every 60 days on average, and the other closer to the findings of Hu and Geng [73], with reinfection every 200 days. These checks help validate our findings on the maximum daily number of infections.

As a key predictor in our study, we adjust the disease’s degree of infectiousness by setting the probability that an exposed individual will infect one of their contacts. Specifically, infectiousness is moderated through a wide range, approximating differing levels of social distancing, masking, and other COVID-aware behaviors, as well as the base transmissibility of the disease. This probability for each pair of contacts is further weighted by their level of interaction. To reflect to the variability of infectiousness given different COVID-19 variants and different diseases, the range of infectiousness, *β*_*se*_, is set from 0.01 to 0.25, indicating that for a unit level of contact, the expected time for one contact to infect another person ranges from 100 days to 4 days. To contextualize this parameter within our network model, considering that the average node degree in each final network is approximately 2.4, if all contacts of an average individual in the network were infected, the expected time until infection for that individual would range from 41.67 days to 1.67 days.

Studies have has shown that the average incubation period for COVID-19 variants ranges from 5.0 days for the Alpha variant to 3.42 for the Omicron Variant [74]. To cover this variation, we set our exposed to infection parameter, *β*_*ei*_, to 1/2 or 1/5, corresponding to an incubation period of 2 days and 5 days. To account for the recovery rate, we set *β*_*ei*_ at 0.1, an expected time to recovery at 10 days, which lies in the middle of recovery durations listed by George et. al [75].

All parameters tested in the simulation are documented in Table 1. Overall, the parameters chosen in this study are meant to constitute an illustration of a more general phenomenon. Determining what extent our findings generalize across every choice of parameters constitutes important future work.

**Table 1.** Description and Notation for Parameters – Including Ranges Employed and Example Descriptions of How These Translate to Our Simulation of Disease Spread.

| Parameter | Description | Value |
| --- | --- | --- |
| $\beta_{se}$ | The rate at which the infected expose their contact partners | [0.01, 0.02, 0.05, 0.1, 0.2, 0.25]<br>0.10 corresponds to once in 10 days, per contact weight with infected individuals |
| $\beta_{ei}$ | The rate at which exposed individuals become infectious | 1/2, 1/5 (e.g., 1/5 corresponds to once in 5 days, on average) |
| $\beta_{ir}$ | The rate at which infected individuals recover from infection | 1/10 (corresponds to once in 10 days, on average) |
| $\beta_{rs}$ | The rate at which recovered individuals become susceptible again | [0, 1/60, 1/200] (e.g., 0 corresponds to never, 1/60 corresponds to once in 60 days on average) |
| $inf_0$ | At the start of the simulation, a constant percentage of random individuals are infected | 5% |
| $\alpha$ | The skewness of the network and the presence of hubs | [-4, -2, 0, 1, 1.5, 2, 2.5, 3, 3.2, 3.5, 3.8, 4]<br>0 means no skewness comparing to the base network, while a positive value means the network degree distribution is more skewed, indicating the presence of hubs |

## RESULTS

Our simulation models offer novel insights into the relationship between the degree distribution of social networks and the spread of infectious diseases. For this study, we kept *β*_*ir*_ fixed at 0.1 and *β*_*rs*_ at 0, and randomly create 300 base networks with 10,000 individuals for each unique set of epidemic-related parameters (i.e., *β*_*se*_, *β*_*ei*,_ *β*_*ir*_ and *inf*_0_). On each base network, we randomly generated 12 final networks via the preferential attachment process where each of them corresponds to one skewness parameter. We run one simulation on each of the final network generated, which results in 300 simulations for each skewness and epidemic parameter combinations. We ran each simulation until the system reaches equilibrium: in an SEIR model, this means all individuals are recovered and no more individuals are infected.

We first explore final size of an outbreak: The proportion of people having been infected at the end of a single outbreak given varied disease infectiousness (*β*_*se*_), network degree skewness, and exposure to infection rate, using an SEIR model. Figure 4 shows a key metric concerning the final size of a single outbreak of simulated epidemic: The numerical difference of final size relative to the zero-skewness network, given *β*_*ei*_ = 0.2.

**Fig 4.**
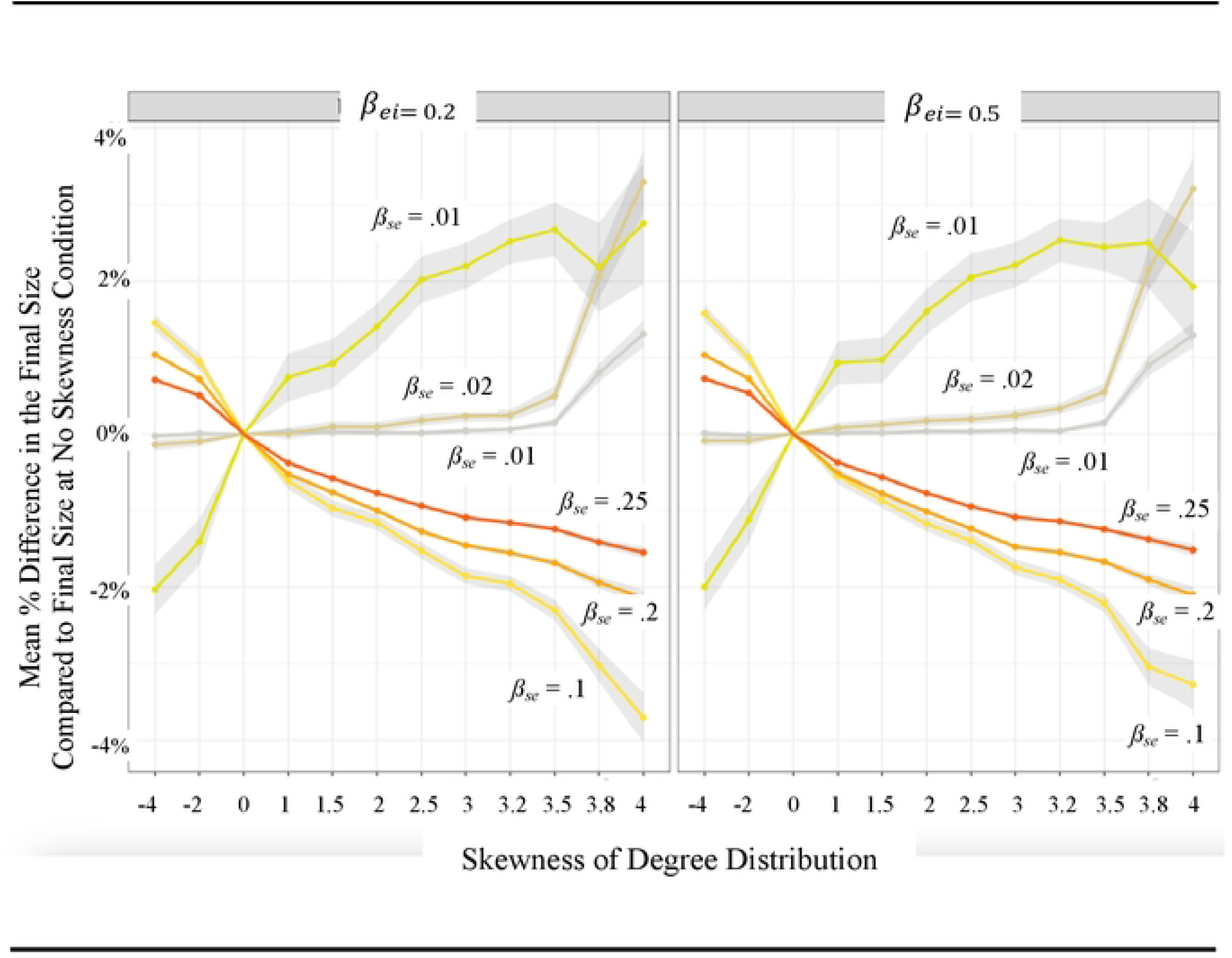
Impact of the Presence of Hubs/Skewness in Social Contact Networks on Final Epidemic Size in Communities. Figure 4 demonstrates the influence of degree-skewness, and thus the prevalence of hubs, in casual social contact networks on the total number of uninfected individuals. This figure takes into account the varied infectiousness of the disease (*β*_*se*_), network skewness, and exposure to infectious rate (*β*_*ei*_). In these simulations, *β*_*ir*_ = 0.1, *β*_*rs*_ = 0 for each parameter combination. The axis presents the average numerical difference in final sizes between a foci-network with degree-skewness and one with zero skewness given the same base network. 95% confidence bands are included. For each contagion rate, 300 base foci-networks were simulated. These base networks were then modified by varying preferential attachment rules, guided by different skewness parameters. Each modified network comprises 10,000 individuals.

In our simulations, while keeping other transmission parameters stable, we found that higher contagion rates (*β*_*se*_ = 0.1,0.2,0.25), networks that have a more skewed degree distribution than the less-skewed ones tend to result in fewer individuals infected in total, assuming the overall number of contacts within the population remains the same. Conversely, at lower contagion rates (*β*_*se*_ = 0.1,0.2,0.25), more individuals in total are infected in networks with higher degree skewness than those with lower degree skewness. For instance, with a contagion rate of *β*_se_ = 0.02 and *β*_se_ = 0.2, high skewness exposes more individuals to risk compared to a zero-skewness condition: foci-networks with *α* = 4 protects 329 (3.3% of the total population) individuals fewer on average than its zero-skewness counterparts built on the same base networks. Conversely, at *β*_se_ = 0.01 and *β*_se_ = 0.2, high skewness results in greater protection than its zero-skewness counterpart: foci-networks with *α* = 4 protects 370 (3.7% of the total population) individuals more on average than its zero-skewness counterparts built on the same base networks.

In addition, the effect of degree skewness is strongest when the disease is neither extremely contagious nor very mild. This could be found by comparing the absolute difference between the number of total infected individuals in a highly-skewed network versus a zero-skewness network (*α* = 0) on different contagion rates (Figure 4). Again, at *β*_se_ = 0.2, a highly skewed network (*α* = 4) protects on average 3.7% more people in the population from being infected given than a zero-skewness network with a relatively high contagion rate of *β*_se_ = 0.1, while it only protects an average of 1.5% more individuals than a zero-skewness network when the contagion rate is even higher at *β*_se_ = 0.25. Table 2 details the proportion of the total population that have been infected at the conclusion of the simulated epidemic without re-infection. While the interpretation uses simulations with *β*_*ei*_ = 0.2, conclusions are similar at *β*_*ei*_ = 0.5.

**Table 2.**
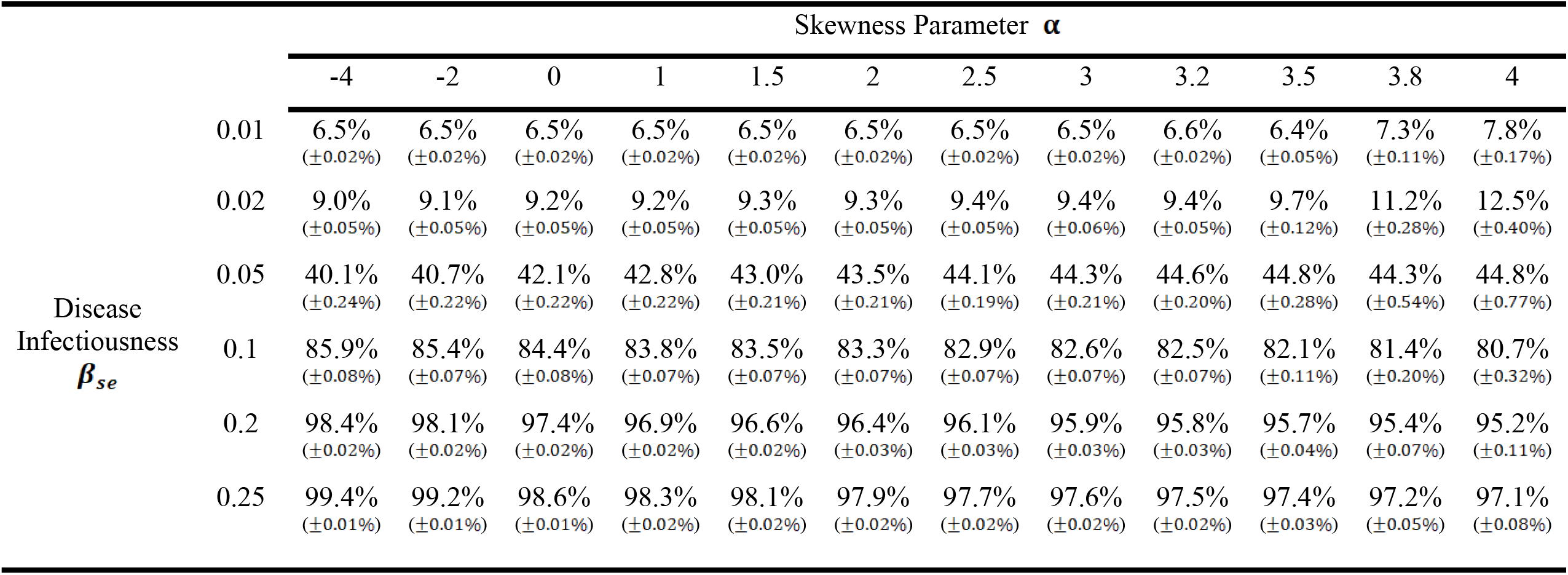
Average Final Size (i.e. Proportion of Total Population Infected at the Conclusion of an Epidemic), by the infectiousness of the disease (*β*_*se*_) and the Presence of Hubs (Network Skewness)

In substantive terms, these findings challenge conventional wisdom about the role of hubs – or highly connected individuals – in disease transmission. Specifically, when the total level of contact within a population is held constant, though hubs themselves are connected to a greater number of individuals, their presence also results in a median individual who is less well-connected, which may form a demographic that is potentially resistant to infection. Our results indicate that infectiousness of the disease influences whether a skewed network structure protects the community or exposes the community to higher risk, highlighting the importance of tailored public health strategies.

While the overall number of cases during an outbreak is important, the number of individuals being infected at the peak of the outbreak offers a unique and critical insight. This number serves as a proxy for the peak demand on healthcare resources, such as hospital beds and medical supplies. Figure 5 presents the proportion of infected individuals at the peak of infection, *β*_*ei*_ = 0.2. When we keep the infectiousness of the disease steady, networks with more pronounced hub structures –indicated by higher skewness – are linked to a surge in peak infections, particularly when the disease has moderate infectiousness (*β*_*se*_ = 0.02,0.05,0.1). However, when a disease is highly infectious, the impact of network skewness appears to diminish: there is not many substantive differences by network skewness in the maximum number of individuals being infected at the same time. Corresponding point estimates are found in Tables 3 and 4. To validate these findings, robustness tests were conducted using SEIRS models with 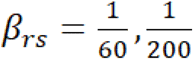. These tests, simulating for 500 days with 300 replications for each parameter set, yield similar conclusion and are detailed in Appendix Figure A1.

**Table 3.** Average Infected Peak Prevalence, Varied by the Infectiousness of the Disease (*β*_*se*_) and Presence of Hubs (Network Skewness)

| | | Skewness Parameter $\alpha$ | | | | | | | | | | | |
| --- | --- | --- | --- | --- | --- | --- | --- | --- | --- | --- | --- | --- | --- |
|  |  | -4 | -2 | 0 | 1 | 1.5 | 2 | 2.5 | 3 | 3.2 | 3.5 | 3.8 | 4 |
| Disease<br>Infectiousness<br>$\beta_{se}$ | 0.01 | 5.0%<br>($\pm 0.00\%$) | 5.0%<br>($\pm 0.00\%$) | 5.0%<br>($\pm 0.00\%$) | 5.0%<br>($\pm 0.00\%$) | 5.0%<br>($\pm 0.00\%$) | 5.0%<br>($\pm 0.00\%$) | 5.0%<br>($\pm 0.00\%$) | 5.0%<br>($\pm 0.00\%$) | 5.0%<br>($\pm 0.00\%$) | 5.0%<br>($\pm 0.00\%$) | 5.0%<br>($\pm 0.00\%$) | 5.0%<br>($\pm 0.00\%$) |
| | 0.02 | 5.0%<br>($\pm 0.00\%$) | 5.0%<br>($\pm 0.00\%$) | 5.0%<br>($\pm 0.00\%$) | 5.0%<br>($\pm 0.00\%$) | 5.0%<br>($\pm 0.00\%$) | 5.0%<br>($\pm 0.00\%$) | 5.0%<br>($\pm 0.00\%$) | 5.0%<br>($\pm 0.00\%$) | 5.0%<br>($\pm 0.00\%$) | 5.0%<br>($\pm 0.00\%$) | 5.0%<br>($\pm 0.00\%$) | 5.0%<br>($\pm 0.00\%$) |
| | 0.05 | 5.0%<br>($\pm 0.00\%$) | 5.0%<br>($\pm 0.00\%$) | 5.0%<br>($\pm 0.00\%$) | 5.0%<br>($\pm 0.00\%$) | 5.0%<br>($\pm 0.00\%$) | 5.0%<br>($\pm 0.00\%$) | 5.0%<br>($\pm 0.00\%$) | 5.0%<br>($\pm 0.00\%$) | 5.0%<br>($\pm 0.01\%$) | 5.2%<br>($\pm 0.06\%$) | 5.6%<br>($\pm 0.15\%$) | 6.8%<br>($\pm 0.22\%$) |
| | 0.1 | 14.8%<br>($\pm 0.06\%$) | 14.8%<br>($\pm 0.06\%$) | 15.1%<br>($\pm 0.06\%$) | 15.2%<br>($\pm 0.06\%$) | 15.2%<br>($\pm 0.06\%$) | 15.3%<br>($\pm 0.06\%$) | 15.4%<br>($\pm 0.06\%$) | 15.4%<br>($\pm 0.06\%$) | 15.5%<br>($\pm 0.06\%$) | 15.6%<br>($\pm 0.09\%$) | 16.1%<br>($\pm 0.18\%$) | 16.4%<br>($\pm 0.28\%$) |
| | 0.2 | 29.0%<br>($\pm 0.06\%$) | 28.9%<br>($\pm 0.06\%$) | 28.8%<br>($\pm 0.06\%$) | 28.7%<br>($\pm 0.06\%$) | 28.7%<br>($\pm 0.06\%$) | 28.6%<br>($\pm 0.06\%$) | 28.6%<br>($\pm 0.06\%$) | 28.6%<br>($\pm 0.06\%$) | 28.5%<br>($\pm 0.06\%$) | 28.6%<br>($\pm 0.07\%$) | 28.9%<br>($\pm 0.15\%$) | 29.0%<br>($\pm 0.21\%$) |
| | 0.25 | 32.9%<br>($\pm 0.06\%$) | 32.7%<br>($\pm 0.06\%$) | 32.5%<br>($\pm 0.06\%$) | 32.4%<br>($\pm 0.07\%$) | 32.3%<br>($\pm 0.06\%$) | 32.3%<br>($\pm 0.06\%$) | 32.2%<br>($\pm 0.06\%$) | 32.2%<br>($\pm 0.06\%$) | 32.2%<br>($\pm 0.06\%$) | 32.1%<br>($\pm 0.07\%$) | 32.4%<br>($\pm 0.14\%$) | 32.7%<br>($\pm 0.19\%$) |

*Note:* In this simulation, $\beta_{ei} = 0.2$ (exposure to infection rate). $\beta_{ir} = 0.1$ , $\beta_{rs} = 0$ . For each parameter combination, 300 simulations were run.

**Table 4.** Average Infected Peak Prevalence, Varied by the Infectiousness of the Disease (*β*_*se*_) and Presence of Hubs (Network Skewness)

| | | Skewness Parameter $\alpha$ | | | | | | | | | | | |
| --- | --- | --- | --- | --- | --- | --- | --- | --- | --- | --- | --- | --- | --- |
|  |  | -4 | -2 | 0 | 1 | 1.5 | 2 | 2.5 | 3 | 3.2 | 3.5 | 3.8 | 4 |
| Disease<br>Infectiousness<br>$\beta_{se}$ | 0.01 | 5.0%<br>($\pm 0.00\%$) | 5.0%<br>($\pm 0.00\%$) | 5.0%<br>($\pm 0.00\%$) | 5.0%<br>($\pm 0.00\%$) | 5.0%<br>($\pm 0.00\%$) | 5.0%<br>($\pm 0.00\%$) | 5.0%<br>($\pm 0.00\%$) | 5.0%<br>($\pm 0.00\%$) | 5.0%<br>($\pm 0.00\%$) | 5.0%<br>($\pm 0.00\%$) | 5.0%<br>($\pm 0.00\%$) | 5.0%<br>($\pm 0.00\%$) |
| | 0.02 | 5.0%<br>($\pm 0.00\%$) | 5.0%<br>($\pm 0.00\%$) | 5.0%<br>($\pm 0.00\%$) | 5.0%<br>($\pm 0.00\%$) | 5.0%<br>($\pm 0.00\%$) | 5.0%<br>($\pm 0.00\%$) | 5.0%<br>($\pm 0.00\%$) | 5.0%<br>($\pm 0.00\%$) | 5.0%<br>($\pm 0.00\%$) | 5.0%<br>($\pm 0.00\%$) | 5.0%<br>($\pm 0.00\%$) | 5.0%<br>($\pm 0.00\%$) |
| | 0.05 | 5.1%<br>($\pm 0.02\%$) | 5.1%<br>($\pm 0.02\%$) | 5.2%<br>($\pm 0.03\%$) | 5.3%<br>($\pm 0.03\%$) | 5.3%<br>($\pm 0.03\%$) | 5.4%<br>($\pm 0.04\%$) | 5.4%<br>($\pm 0.04\%$) | 5.5%<br>($\pm 0.04\%$) | 5.6%<br>($\pm 0.05\%$) | 5.7%<br>($\pm 0.07\%$) | 7.1%<br>($\pm 0.21\%$) | 7.9%<br>($\pm 0.26\%$) |
| | 0.1 | 18.8%<br>($\pm 0.07\%$) | 18.9%<br>($\pm 0.07\%$) | 19.0%<br>($\pm 0.07\%$) | 19.2%<br>($\pm 0.07\%$) | 19.3%<br>($\pm 0.07\%$) | 19.5%<br>($\pm 0.07\%$) | 19.5%<br>($\pm 0.07\%$) | 19.6%<br>($\pm 0.07\%$) | 19.7%<br>($\pm 0.07\%$) | 19.7%<br>($\pm 0.10\%$) | 20.2%<br>($\pm 0.22\%$) | 20.9%<br>($\pm 0.31\%$) |
| | 0.2 | 37.3%<br>($\pm 0.07\%$) | 37.2%<br>($\pm 0.07\%$) | 37.0%<br>($\pm 0.07\%$) | 36.9%<br>($\pm 0.06\%$) | 36.9%<br>($\pm 0.07\%$) | 36.8%<br>($\pm 0.07\%$) | 36.8%<br>($\pm 0.06\%$) | 36.8%<br>($\pm 0.07\%$) | 36.8%<br>($\pm 0.07\%$) | 36.8%<br>($\pm 0.08\%$) | 37.1%<br>($\pm 0.17\%$) | 37.4%<br>($\pm 0.24\%$) |
| | 0.25 | 42.5%<br>($\pm 0.07\%$) | 42.4%<br>($\pm 0.07\%$) | 42.1%<br>($\pm 0.07\%$) | 42.0%<br>($\pm 0.06\%$) | 41.9%<br>($\pm 0.07\%$) | 41.8%<br>($\pm 0.07\%$) | 41.6%<br>($\pm 0.06\%$) | 41.6%<br>($\pm 0.06\%$) | 41.6%<br>($\pm 0.06\%$) | 41.6%<br>($\pm 0.08\%$) | 42.0%<br>($\pm 0.17\%$) | 42.1%<br>($\pm 0.21\%$) |

*Note:* In this simulation, $\beta_{ei} = 0.5$ (exposure to infection rate). $\beta_{ir} = 0.1$ , $\beta_{rs} = 0$ . For each parameter combination, 300 simulations were run.

**Fig 5.**
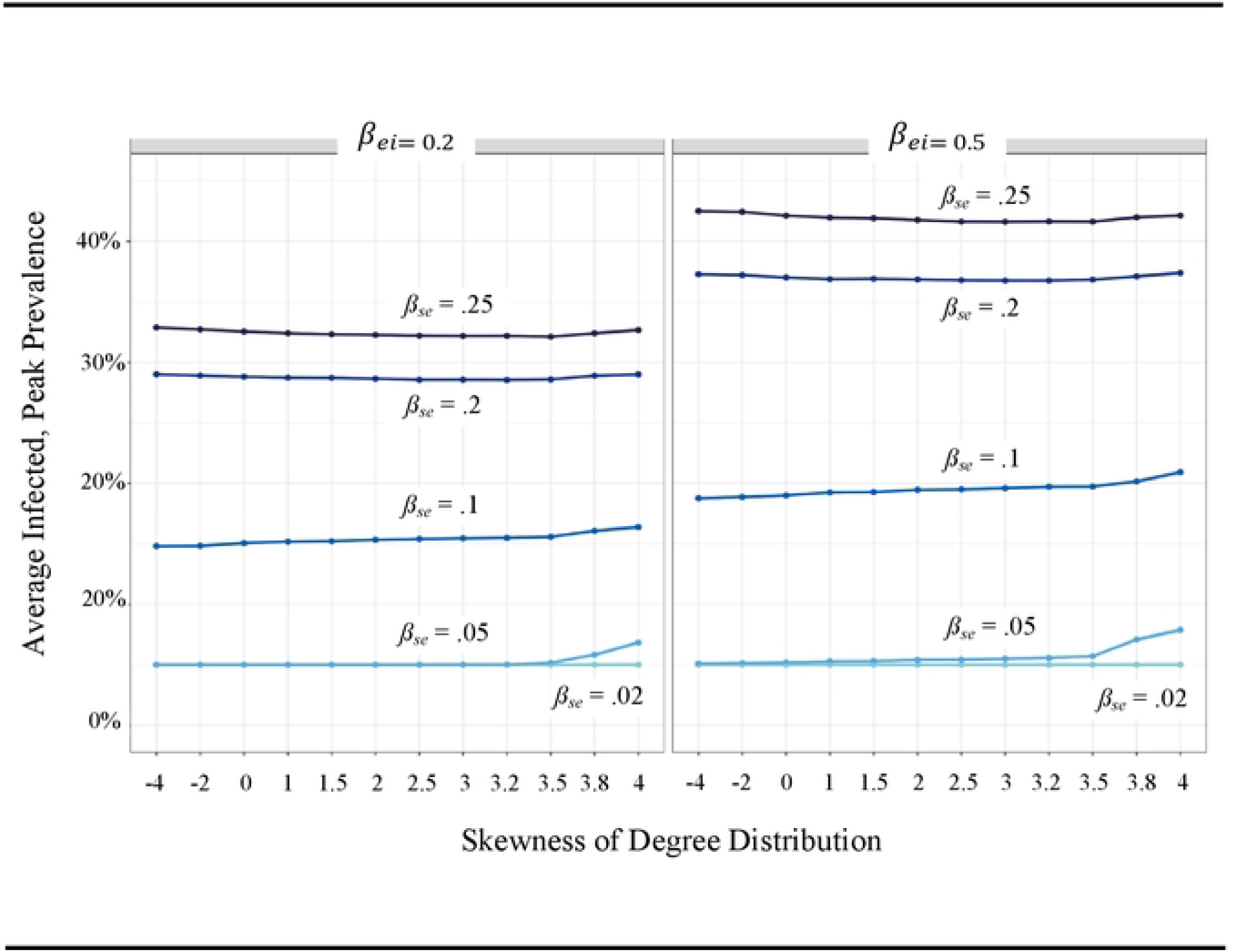
Impact of the Presence of Hubs/Skewness in Casual Contact Networks on Peak Prevalence of Infection in Communities by Disease infectiousness (*ß*_se_) *Note:* Estimates are varied by the infectiousness of the disease (*β*_*se*_), network skewness and exposure to infectious rate (*β*_*ei*_). In this simulation, *β*_*ir*_ = 0.1, *β*_*rs*_ = 0. For each parameter combination, 300 simulations were run, and 95% confidence bands are drawn.

## DISCUSSION

Health and mortality are indelibly linked to social structure, for individuals and for the broader community. We have begun to understand the importance that social networks play as a part of the social-structural underpinnings of numerous health outcomes [5, 6, 7, 8, 9, 10, 11, 12, 13, 14]. As it pertains to the subject of this paper, there is a rapidly growing body of evidence showing that social networks shape the incidence of diseases within communities, and their spread across communities [1, 2, 3, 4]. With the recent emergence and combination of numerous disease epidemics, scholars have begun to pay more attention to the role that larger social network structure plays in shaping disease outbreaks and spread. We build on recent work in this vein to explore how social network structure might affect the spread of disease throughout the community.

## CONCLUSIONS

Our findings suggest that networks that have more skewed degree distributions lead to greater risk of epidemic spread overall, as has been documented in numerous other papers [31, 32, 76]. We expand on this work by showing that this effect depends on disease transmissibility. Our work shows that the network-structural effect decreases as disease transmissibility increases.

There are several interpretations that are consistent with these findings. First, we confirm empirical and simulation evidence [59, 64] that epidemics spread via hubs, but this effect depends on the disease’s infectiousness or transmissibility. It is possible that hubs themselves are disproportionately affected by more infectious diseases: Hubs, or “superspreaders,” as they have been called, have more exposure to people who are infected early on, and therefore are more likely to isolate early on. One recent study [77] therefore cautioned that so-called superspreaders should also be viewed as “super-receivers.” High transmissibility can take such hubs out of the network, and thus leave the network disproportionately unconnected at an earlier stage in an epidemic.

An equally plausible interpretation is that any social structural feature –social network structure, social resources, or other factors – simply becomes less important as diseases become more virulent. In such scenarios, networks or other social factors do not matter; infection and spread happens due to the sheer transmissibility of the disease. A highly transmissible disease is a disease that defies social structure and just runs rampant over it.

This should not detract us from exploring further the role that social networks, structure and heterogeneity play in shaping the epidemic spread of disease. As mentioned earlier, there is much research on how having a large number of social connections affects the spread of disease and the overall risk of epidemics. In this case, it appears that this empirical account will be more relevant when one considers less transmissible diseases or less contagious variants of diseases like COVID-19.

These findings have potential implications for the epidemic spread of diseases in certain social contexts. From a social networks perspective, these results confirm that a highly heterogeneously connected population is at greater risk for epidemic spread, especially with a relatively slowly spreading virus. Social distancing, vaccination, quarantining, and other mitigation measures can reduce the risk of epidemic spread – but these measures are particularly useful at this stage where there are numerous hubs, stars, or centers where diseases can be easily transmitted to numerous people at the same time. It is at this point that the need for public policy measures is most urgent.

### Limitations and Directions

There are several limitations to this study that call for future study. For one, although maintaining a constant overall contact structure in our simulations is crucial (as this significantly influences the spread of disease), this approach may overlook the relationship between degree distribution skewness and overall contact levels. In real-world networks, total contact rate can correlate with degree distribution skewness due to factors like population characteristics and occupational distributions, so it is important to note that our findings do not take in consideration cases where both total contact and degree distribution skewness increase simultaneously.

Additionally, our results are specific to the method used to generate the network. Discrepancies may arise when compared to other network generation methods with similar overall contact and degree distributions. Future research should evaluate our findings using different network generation methods.

We did not test how different model formulations affect the impact of social network structure on spread. Evidence suggests that the epidemic spread of disease can only partially capture models of spread by estimating *R*_0_, using SIR or SEIR models alone, as it also requires taking into account the existence of asymptomatic, pre-symptomatic, mild, and severe categories of infection [65]. Future research should consider these different scenarios as potential conditions on the role that social network structure plays in the epidemic spread of disease.

Our simulations, while informed by empirical data, lack primary data on network degree distribution and variant transmissibility. Future research should track both social network structure and disease spread within a community over time, and the relationship that network change (e.g., due to personal- and community-level network change play in shaping individuals’ susceptibility to disease and communities’ abilities to organize social support, prevention, and treatment efforts [66]. However, assuming that the data upon which we build our models are reliable, we believe our models offer valuable insights into the interplay between social network degree distribution, disease transmissibility, and epidemic spread. Expanding on this work should aide the scientific community in informing public health responses in the advent of current and future disease outbreaks, and more generally provide us with a better sense of how health is rooted in social behavior and structure.

## Data Availability

The authors conducted simulations. No real-life data were used in developing this manuscript.

## ACKNOWLEDGEMENTS

This project was funded by the National Science Foundation, Division of Mathematical Sciences (Award #2230023).

This project benefited from the comments and suggestions of Erin York Cornwell, Edward O. Laumann, Gen Meredith, Peter Frazier, Varun Gande, and other members from the NSF Award research group. We also thank Camron Chappel for research assistance.

## REFERENCES

1 Ehsan A, Klaas HS, Bastianen A, Spini D. Social capital and health: A systematic review of systematic reviews. SSM. Popul. Health., 2019; 8 100425.

2 Kawachi I, Kennedy BP, Glass R. Social capital and self-rated Health: A contextual analysis. Am. J. Public Health., 1999; 89 1187–1193.

3 Poortinga W. Social capital: An individual or collective resource for health? Soc. Sci. Med., 2006; 62 292–302.

4 Rodgers J, Valuev VA, Hswen Y, Subramanian SV. Social capital and physical health: An updated review of the literature for 2007-2018. Soc. Sci. Med., 2019; 236 112360.

5 Berkman LF, Glass T, Brissette I, Seeman TE. From social integration to health: Durkheim in the new millennium. Soc. Sci. Med., 2000; 51 843–857.

6 Carr D, Umberson D. The social psychology of stress, health, and coping. In: DeLamater J, Ward A, editors. Handbook of Social Psychology. Dordrecht: Springer Netherlands; 2013. pp. 465–487.

7 De Silva MJ, McKenzie K, Harpham T, Huttly SR. Social capital and mental illness: A systematic review. J. Epidemiol. Community Health., 2005; 59 619–627.

8 Kawachi I, Subramanian SV, Kim D, editors. Social capital and health. New York: Springer; 2008.

9 Smith KP, Christakis NA. Social networks and health. Annu. Rev. Sociol., 2008; 34 405–429.

10 Thoits PA. Mechanisms linking social ties and support to physical and mental health. J. Health Soc. Behav., 2011; 52 145–161.

11 Thomas PA, Liu H, Umberson D. Family relationships and well-being. Innov. Aging, 2017; 1 1–11.

12 Turner B. Social capital, inequality and health: The Durkheimian revival. Soc. Theory Health, 2003; 1 4–20.

13 York Cornwell E, Waite LJ. Social disconnectedness, perceived isolation, and health among older adults. J. Health Soc. Behav., 2009; 50 31–48.

14 Zhang JD, Centola. Social networks and health: New developments in diffusion, online and offline. Annu, Rev., Sociol., 2019; 45 91–109.

15 Szreter S, Woolcock M. Health by association? Social capital, social theory, and the political economy of public health. Int. J. Epidemiol., 2004; 33 650–667.

16 Chen S, Prettner K, Kuhn M, et al. Climate and the spread of COVID-19. Sci. Rep., 2002; 11(1). doi:10.1038/s41598-021-87692-z

17 Kadi N, Khelfaoui M. Population density: A factor in the spread of COVID-19 in Algeria: Statistic study. Bull. Nat. Res. Cent., 2020; 44(1). doi:10.1186/s42269-020-00393-x

18 Fe Feinhandler I, Cilento B, Beauvais B, et al. Predictors of death rate during the COVID-19 pandemic. Healthcare., 2020; 8(3) 339. doi:10.3390/healthcare8030339

19 Ganasegeran K, FadzlyM, Jamil A, et al. Influence of population density for COVID-19 spread in Malaysia: An ecological study. Int. J. Environ. Res. Public Health., 2021; 18(18) 9866. doi:10.3390/ijerph18189866

20 Lai L, Yan K, Webster C, et al. The nature of cities and the COVID-19 pandemic. Curr. Opin. Environ. Sustain., 2020; 46 27–31. doi:10.1016/j.cosust.2020.08.008

21 Laumann EO, Youm Y. Racial/ethnic group differences in the prevalence of sexually transmitted diseases in the United States: A network explanation. Sex. Trans. Dis., 1999; 26(5) 250–261. doi:10.1097/00007435-199905000-00003

22 Malik AA, McFadden SAM, Elharake J, Omer SB. Determinants of COVID-19 vaccine acceptance in the U.S. EClinicalMedicine. 2020; 26 100495. doi:10.1016/j.eclinm.2020.100495

23 Youm Y, Laumann EO. Social network effects on the transmission of sexually transmitted diseases. Sex. Transm. Dis., 2002; 29 689–697.

24 Yum S. Social network analysis for coronavirus (COVID-19) in the United States. Soc. Sci. Q., 2020; 101(4) 1642–1647. doi:10.1111/ssqu.12808

25 Bansal S, Grenfell BT, Meyers LA. When individual behaviour matters: Homogeneous and network models in epidemiology. J. R. Soc. Interface., 2007; 4(16) 879–891. doi:10.1098/rsif.2007.1100

26 Volz EM, Miller JC, Galvani A, Ancel Meyers L. Effects of heterogeneous and clustered contact patterns on infectious disease dynamics. PLoS Comput. Biol., 2011; 7(6). doi:10.1371/journal.pcbi.1002042

27 Fujimoto K, Flash CA, Kuhns LM, et al. Social networks as drivers of syphilis and HIV infection among young men who have sex with men. Sex. Transm. Infect., 2008; 94(5) 365–371. doi:10.1136/sextrans-2017-053288

28 Shah NS, Iveniuk J, Muth SQ, et al. Structural bridging network position is associated with HIV status in a younger black men who have sex with men epidemic. AIDS Behav., 2013; 18(2) 335–345. doi:10.1007/s10461-013-0677-8

29 Liotta G, Marazzi MC, Orlando S, Palombi L. Is social connectedness a risk factor for the spreading of COVID-19 among older adults? The Italian paradox. PloS ONE. 2020; 15(5). doi:10.1371/journal.pone.0233329

30 Marlow T, Makovi K, Abrahao B. Neighborhood isolation during the COVID-19 pandemic. Soc. Sci., 2021; 8 170–190. doi:10.15195/v8.a9

31 Eguiluz VM, Klemm K. Epidemic threshold in structured scale-free networks. Phys. Rev. Lett., 2002; 89(10). doi:10.1103/physrevlett.89.108701

32 Kleczkowski A, Grenfell BT. Mean-field-type equations for spread of epidemics: The ‘small world’ model. Physica A., 1999; 274(1-2) 355–360. doi:10.1016/s0378-4371(99)00393-3

33 Barthélemy M, Barrat A, Pastor-Satorras R, Vespignani A. Dynamical patterns of epidemic outbreaks in complex heterogeneous networks. J. Theor. Biol., 2005; 235(2) 275–288. doi:10.1016/j.jtbi.2005.01.011

34 Hindes J, Schwartz IB. Epidemic extinction and control in heterogeneous networks. Phys. Rev. E., 2016; 117(2) 028302. doi:10.1103/physrevlett.117.028302

35 Jo W, Chang D, You M, Ghim G-H. A social network analysis of the spread of COVID-19 in South Korea and policy implications. Sci. Rep., 2021; 11(1). doi:10.1038/s41598-021-87837-0

36 Block P, Hoffman M, Raabe IJ, et al. Social network-based distancing strategies to flatten the COVID-19 curve in a post-lockdown world. Nat. Hum. Behav., 2020; 4(6) 588–596. doi:10.1038/s41562-020-0898-6

37 Glass RJ, Glass LM, Beyeler WE, Min HJ. Targeted social distancing designs for pandemic influenza. Emerg. Infect. Dis., 2006; 12(11) 1671–1681. doi:10.3201/eid1211.060255

38 Makridis CA, Wu C. Correction: How social capital helps communities weather the COVID-19 pandemic. PloS ONE. 2021; 16(9) e0245135. doi:10.1371/journal.pone.0258021

39 Schlosser F, Maier BF, Jack O, et al. COVID-19 lockdown induces disease-mitigating structural changes in mobility networks. Proc. Natl. Acad. Sci., U. S. A., 2020; 117(52) 32883–32890. doi:10.1073/pnas.2012326117

40 Sjödin H, Wilder-Smith A, Osman S, et al. Only strict quarantine measures can curb the coronavirus disease (COVID-19) outbreak in Italy. Euro. Surveill., 2020; 25(13) 2000280. doi:10.2807/1560-7917.es.2020.25.13.2000280

41 Stockmaier S, Stroeymeyt N, Shattuck EC, et al. Infectious diseases and social distancing in nature. Sci., 2021; 371 (6533). doi:10.1126/science.abc8881

42 Weeden K, Cornwell B. The small-world network of college classes: Implications for epidemic spread on a university campus. Soc. Sci., 2020; 7 222–241. doi:10.15195/v7.a9

43 Weeden K, Cornwell B, Park B. Still a small world? University course enrollment networks before and during the COVID-19 pandemic. Soc. Sci., 2021; 8 73–82.

44 Britton T, Ball F, Trapman P. A mathematical model reveals the influence of population heterogeneity on herd immunity to SARS-COV-2. Sci., 2020; 369(6505) 846–849. doi:10.1126/science.abc6810

45 Liu Y, Gu Z, Liu J. Uncovering transmission patterns of COVID-19 outbreaks: A region-wide comprehensive retrospective study in Hong Kong. EClinicalMedicine. 2021; 36 100929. doi:10.1016/j.eclinm.2021.100929

46 Althouse BM, Wenger EA, Miller JC, et al. Superspreading events in the transmission dynamics of SARS-CoV-2: Opportunities for interventions and control. PloS Biol., 2020; 18(11) doi:10.1371/journal.pbio.3000897.

47 Karaivanov A. A social network model of COVID-19. PloS ONE., 2020; 15(10) e0240878. doi:10.1371/journal.pone.0240878

48 Kuchler T, Russel D, Stroebel J. JUE insight: The geographic spread of COVID-19 Correlates with the structure of social networks as measured by Facebook. J. Urban Econ., 2022; 127 103314.

49 Liu Y, Gu Z, Liu J. Uncovering transmission patterns of COVID-19 outbreaks: A region-wide comprehensive retrospective study in Hong Kong. EClinicalMedicine. 2021; 36 100929. doi:10.1016/j.eclinm.2021.100929

50 Liu Y, Rocklöv J. The reproductive number of the delta variant of SARS-COV-2 is far higher compared to the ancestral SARS-COV-2 virus. J. Travel Med., 2021; 28(7). doi:10.1093/jtm/taab124

51 Sheikhi F, Yousefian N, Nehranipoor P, Kowsari Z. Estimation of the basic reproduction number of alpha and delta variants of COVID-19 pandemic in Iran. PloS ONE. 2022; 17(5) 0265489. doi:10.1371/journal.pone.0265489

52 Guerra FM, Bolotin S, Lim G, et al. The basic reproduction number (R0) of measles: A systematic review. Lancet Infect. Dis., 2017; 17(12) e420–e428. doi:10.1016/s1473-3099(17)30307-9

53 Feld SL. The focused organization of social ties. Am. J. Sociol., 1981; 86(5) 1015–1035.

54 Garnett GP, Anderson RM. Contact tracing and the estimation of sexual mixing patterns. J. Sex. Transm. Dis., 1993; 20(4)181-191. doi:10.1097/00007435-199307000-00001

55 Muchnik L, Pei S, Parra LC, et al. Origins of power-law degree distribution in the heterogeneity of human activity in social networks. Sci. Rep., 2013; 3(1). doi:10.1038/srep01783

56 Newman ME. Models of the small world. J. Stat. Phys., 2000; 101(3/4) 819-841. doi:10.1023/a:1026485807148

57 Barabási A-L, Albert R. Emergence of scaling in random networks. Sci., 1999; 286(5439) 509–512. doi:10.1126/science.286.5439.509

58 Watts DJ, Strogatz SH. Collective dynamics of ‘small-world’ networks. Nat., 1998; 393(6684) 440–442. doi:10.1038/30918

59 Barthélemy MA, Barrat A, Pastor-Satorras R, Vespignani A. Velocity and hierarchical spread of epidemic outbreaks in scale-free networks. Phys. Rev. Lett., 2004; 92(17) 178701.

60 Snijders TA, Nowicki K. Estimation and prediction for stochastic blockmodels for graphs with latent block structure. J. Classif., 1997; 14(1) 75–100. doi:10.1007/s003579900004

61 Erdős PP, Rényi A. On the evolution of random graphs. Publication of the Mathematical Institute of the Hungarian Academy of Sciences, 1960; 5 17–61.

62 Feld SL. The focused organization of social ties. Am. J. Sociol., 1981; 86(5) 1015–1035

63 Nguyen NN, Nguyen YN, Hoang VT, et al. SARS-CoV-2 reinfection and severity of the disease: A systematic review and meta-analysis. J. Viruses, 2023; 15(4) 967. doi:10.3390/v15040967

64 Adam DC, Wu P, Wong JY, et al. 2020. Clustering and superspreading potential of SARS-CoV-2 infections in Hong Kong. Nat Med., 2020; 26(11) 1714–1719.

65 United States Census Bureau. The number of firms and establishments, employment, and annual payroll by state, industry, and enterprise employment size: 2020. 2020 Statistics of U.S. Businesses. 2020. Available from: https://www.census.gov/programs-surveys/susb.html

66 United States Department of Justice, Office of Juvenile Justice and Delinquency Prevention. Juvenile Population. OJJDP Statistics Briefing Book, 2021. 2021. Available from: https://www.ojjdp.gov/ojstatbb/population/qa01104.asp?qaDate=202.

67 United States Department of Justice, Office of Juvenile Justice and Delinquency Prevention. Juvenile Population. OJJDP Statistics Briefing Book, 2021. 2021. Available from: https://www.ojjdp.gov/ojstatbb/population/qa01104.asp?qaDate=202.

68 United States Department of Agriculture, Economic Research Service. Retail food sales and sales growth. Retail trend, 2023. 2003. Available from: https://www.ers.usda.gov/topics/food-markets-prices/retailing-wholesaling/retail-trends/.

69 Lloyd AL. Realistic distributions of infectious periods in epidemic models: Changing patterns of persistence and dynamics. Theor. Popul. Biol., 2001; 60(1) 59–71.

70 Miller JC, Eon TT. (Epidemics on networks): A fast, flexible Python package for simulation, analytic approximation, and analysis of epidemics on networks. J. Open Source Softw., 2019; 4(44) 1731. doi:10.21105/joss.01731

71 McGee RS, Homburger JR, Williams HE, et al. Model-driven mitigation measures for reopening schools during the COVID-19 pandemic. Proc. Natl. Acad. S. U. S. A., 2021; 118(39) e2108909118. doi:10.1073/pnas.2108909118

72 Guedes AR, Oliveira MS, Tavares BM, et al. Reinfection rate in a cohort of healthcare workers over 2 years of the COVID-19 pandemic. Sci. Rep., 2023; 13(1). doi:10.1038/s41598-022-25908-6

73 Hu G, Geng J. Heterogeneity learning for SIRS model: An application to COVID-19. Stat. Its. Interface., 2021; 14 73–81. doi: 10.48550/arXiv.2007.08047

74 Wu Y, Kang L, Guo Z, et al. Incubation period of COVID-19 caused by unique SARS-COV-2 strains. JAMA Netw. Open., 2022; 5. doi:10.1001/jamanetworkopen.2022.28008

75 George N, Tyagi NK, Prasad JB. COVID-19 Pandemic and its average recovery time in Indian states. Clin. Epidemiol. Glob. Health., 2021; 11 100740. doi:10.1016/j.cegh.2021.100740

76 Newman ME. Spread of epidemic disease on networks. Phys. Rev. E., 2002; 66(1). doi:10.1103/physreve.66.016128

77 Nielsen BF, Simonsen L, Sneppen K. COVID-19 superspreading suggests mitigation by social network modulation. Phys. Rev. Lett., 2021;126(11) 118301. doi:10.1103/physrevlett.126.118301

